# Towards a diagnostic test for sporadic ALS utilising deep learning and SNP microarrays

**DOI:** 10.1101/2025.04.12.25325716

**Authors:** Jiajing Hu, Oliver Pain, Ahmad Al Khleifat, Aleksey Shatunov, Peter M. Andersen, Nazli A. Başak, Johnathan Cooper-Knock, Philippe Corcia, Philippe Couratier, Mamede de Carvalho, Vivian Drory, Marc Gotkine, John E. Landers, Jonathan D. Glass, Russell McLaughlin, Jesús S. Mora Pardina, Karen E. Morrison, Susana Pinto, Monica Povedano, Christopher E. Shaw, Pamela J. Shaw, Vincenzo Silani, Nicola Ticozzi, Philip van Damme, Leonard H. van den Berg, Patrick Vourc’h, Markus Weber, Orla Hardiman, Jan H. Veldink, Project MinE ALS Sequencing Consortium, Richard J.B. Dobson, Alexander Schönhuth, Ammar Al-Chalabi, Alfredo Iacoangeli

**Affiliations:** Maurice Wohl Clinical Neuroscience Institute, Department of Basic and Clinical Neuroscience, Institute of Psychiatry, Psychology and Neuroscience, King’s College London, London, SE5 8AF, UK; Department of Biostatistics and Health Informatics, Institute of Psychiatry, Psychology and Neuroscience, King’s College London, London, SE5 8AF UK; Department of Clinical Science, Umeå University, Umeå SE-901 85, Sweden; Koc University, School of Medicine, Translational Medicine Research Center, NDAL, Istanbul, 34450, Turkey; Sheffield Institute for Translational Neuroscience (SITraN), University of Sheffield, Sheffield S10 2HQ, UK; UMR 1253, Université de Tours, Inserm, Tours 37044, France; Centre de référence sur la SLA, CHU de Tours, Tours 37044, France; Centre de référence sur la SLA, CHRU de Limoges, Limoges, France; UMR 1094, Université de Limoges, Inserm, Limoges 87025, France; Instituto de Fisiologia, Medicine ULisboa for Health, Clinical Research and Innovation, Faculdade de Medicina, Universidade de Lisboa, Lisbon 1649-028, Portugal; Department of Neurology, Tel-Aviv Sourasky Medical Centre, Tel-Aviv 64239, Israel; Faculty of Medical Sciences, Tel-Aviv University 6997801, Israel; Faculty of Medicine, Hebrew University of Jerusalem, Jerusalem 91904, Israel; Agnes Ginges Center for Human Neurogenetics, Department of Neurology, Hadassah Medical Center, Jerusalem 91120, Israel; Department of Neurology, University of Massachusetts Medical School, Worcester, MA 01655, USA; Department of Neurology, Emory University School of Medicine, Atlanta, Georgia, GA 30322, USA; Complex Trait Genomics Laboratory, Smurfit Institute of Genetics, Trinity College Dublin, Dublin D02 PN40, Ireland; ALS Unit, Hospital San Rafael, Madrid, Spain; School of Medicine, Dentistry and Biomedical Sciences, Queen’s University Belfast, Belfast BT9 7BL, UK; Functional Unit of Amyotrophic Lateral Sclerosis (UFELA), Service of Neurology, Bellvitge University Hospital, L’Hospitalet de Llobregat, Barcelona 08907, Spain; Department of Neurology and Laboratory of Neuroscience, Istituto Auxologico Italiano, IRCCS, Milan 20149, Italy; Department of Pathophysiology and Transplantation, “Dino Ferrari” Center, Università degli Studi di Milano, Milan 20122; Department of Neuroscience and Leuven Brain Institute (LBI), KU Leuven, Leuven 3000, Belgium; Department of Neurology, University Hospitals Leuven, Leuven 3000, Belgium; Department of Neurology, UMC Utrecht Brain Center, University Medical Center Utrecht 3584 CX, Netherlands; Service de Biochimie et Biologie molécularie, CHU de Tours, Tours 37044, France; Muskelzentrum/ALS Clinic, HOCH Health Ostschweiz, Kantonsspital St.Gallen, Switzerland; Academic Unit of Neurology, Trinity Biomedical Sciences Institute, Trinity College Dublin, Dublin D02 PN40, Ireland; NIHR Biomedical Research Centre at South London and Maudsley NHS Foundation Trust and King’s College London, UK; Institute of Health Informatics, University College London, London, NW1 2DA, UK; NIHR Biomedical Research Centre at University College London Hospitals NHS Foundation Trust, London, UK; Genome Data Science, Faculty of Technology, Bielefeld University, 33615 Bielefeld, Germany; Department of Clinical Neuroscience, King’s College Hospital, London, SE5 9RS, UK; Perron Institute for Neurological and Translational Science, University of Western Australia Medical School, Perth, Australia

## Abstract

A variety of common and rare genetic factors have been implicated in the development of amyotrophic lateral sclerosis (ALS), and the evidence is that a genetic component is present in most affected individuals. However, our current understanding of ALS genetics causally explains only a small proportion of sporadic cases which represent over 90% of all people with ALS. This limits the utility of genetic testing in screening, diagnosis and management to the 15-20% of people with ALS who carry a known pathogenic variant.

Capsule Networks (CapsNets) constitute a deep learning method that has demonstrated strong performance in using genotyping data to predict individuals at risk for ALS. However, their use is constrained by a lack of generalised, flexible, and validated implementations across comprehensive datasets that account for the technical, biological, and clinical heterogeneity found in real-world disease scenarios. In this study, we build upon this method to address existing limitations, to develop a new model that is validated across diverse ALS populations, can handle discrepancies between genotyping technologies, and is applicable to individual external samples.

Using large-scale datasets from over 47,000 individuals from 13 countries, genotyped with nine different genotyping platforms, our model achieved high precision and sensitivity in distinguishing between individuals with ALS and non-affected controls. Moreover, in simulations of population screening for ALS, its performance was comparable to that of conventional genetic screening for known ALS gene mutations, such as *FUS* and *C9orf72*.

Our results demonstrate that this flexible and validated method could support the development of a genetic screening test for identifying individuals at risk and expediting ALS diagnosis. This would be applicable to all individuals, regardless of their family history or presence of known ALS mutations.

## Background

Amyotrophic lateral sclerosis (ALS) is a devastating disease, characterized by the progressive degeneration of both upper and lower motor neurons, leading to denervation of voluntary muscles and death from respiratory failure [1]. Globally, ALS has an incidence of between 0.6 and 3.8 per 100,000 person-years, a prevalence ranging from 4.1 to 8.4 per 100,000 persons [2, 3] and a lifetime risk of approximately 1 in 300 [4].

There is a substantial genetic component in the development of ALS. Twin-based studies estimate heritability to be ∼60% [5], and mutations in over 40 genes are associated with disease risk or act as disease modifiers [6-11]. Such mutations can explain the majority of affected individuals who exhibit a family history of ALS (FALS), but these represent only 5-10% of people with ALS [1]. When considering all ALS, these gene variants can be found in about 20% of affected people [7, 12]. As a result, the usability of genetic testing in the diagnosis or screening of ALS is limited and the genetics driving ALS in the majority of affected people remains unclear.

To unravel the genetic complexity of ALS, genome-wide association studies (GWASs) have been conducted to identify common genetic markers associated with the disease [13-16]. However, despite their success in identifying common variants associated with ALS risk in several loci, these only represent between 7.2% and 9.5% of ALS heritability, and ALS polygenic risk scores (PRSs) based on such SNPs can explain a limited proportion of genetic variance, ranging from 0.010 to 0.022 [17, 18]. A possible explanation for the discrepancy between the measured heritability and what we can explain with the current known genetic factors, could be insufficient statistical power to discover all important associated variants. However, another contributing factor may be the failure to account for non-linear interactions between genetic variants in additive models utilised for measuring heritability and gene discovery [19].

In recent years, advanced statistical methods and machine learning have been applied in ALS genetic research [20-27]. This includes the development of prognostic models for identifying different risk groups [21], predicting ALS progression and functional impairment [24], and ALS-related gene discovery utilizing functional genomics with GWAS summary statistics [25].

Deep Learning, a subset of machine learning (ML), has demonstrated success in predicting ALS outcomes by leveraging clinical information along with genomic and MRI data [28-30]. Despite these advances, there remains a gap in applying deep learning to genomic data to predict ALS outcomes and occurrence.

Capsule networks (CapsNets) [31], offer a novel deep learning architecture that addresses challenges related to interpretability and extensive training data requirements as they provide improvements in accuracy and reduced data requirements compared to traditional Convolutional Neural Networks (CNNs). One study [32] applied CapsNets to model genome-wide, non-additive gene interactions in large case-control datasets including one comprising thousands of people with ALS and controls, genotyped with SNP microarrays. Their CapsNet outperformed other ML methods and conventional approaches based on PRSs in predicting disease occurrence. Nonetheless, their study primarily focussed on a Dutch cohort which, given the intricate local population structure within the Netherlands, could lead to a performance overestimation [33].

Moreover, the impossibility of utilising the method without reprocessing all samples together from scratch (both training and testing) greatly limits its translational potential and raises uncertainties about its applicability to more intricate and heterogenous datasets involving multiple cohorts, populations, and technologies. In this study we overcome these limitations by constructing a new CapsNet model trained on a multi-cohort dataset, comprising over 47,000 individuals from 13 countries genotyped with 9 different array platforms, that can be used on external samples from different platforms, without the need for reprocessing training and testing data together. Such a flexible and validated model holds potential for setting the basis of the first genetic testing approach based on SNPs that is applicable to all people with ALS.

## Methods

### Data pre-processing, quality controls and imputation for the training and validation data

Genotyping data from 14,791 ALS patients and 26,898 controls across 41 cohorts and 13 countries [34] were used for training and validation (Supplementary Table 1). Standard quality control procedures were followed as detailed in the supplementary material. The 41 cohorts were combined based on genotyping platform and nationality to form 27 case–control strata. Finally, 12,587 cases and 23,503 controls passed quality control (Table 1 and Supplementary Table 2).

**Table 1.** Description of the external dataset cohorts.

|  | Training and validation dataset |  |  | Independent test dataset |  |  |
| --- | --- | --- | --- | --- | --- | --- |
| National cohort | Controls | Cases | Total | Controls | Cases | Total |
| Belgium | 559 | 504 | 1063 | 158 | 246 | 404 |
| Finland | 475 | 516 | 991 | -- | -- | -- |
| France | 1659 | 482 | 2141 | -- | -- | -- |
| Germany | 907 | 1917 | 2824 | -- | -- | -- |
| Ireland | 774 | 572 | 1346 | 234 | 443 | 677 |
| Italy | 1412 | 2388 | 3800 | -- | -- | -- |
| The Netherlands | 7680 | 2116 | 9796 | 887 | 1376 | 2263 |
| Spain | 99 | 126 | 225 | 128 | 273 | 401 |
| Portugal | -- | -- | -- | 13 | 38 | 51 |
| Sweden | 514 | 526 | 1040 | -- | -- | -- |
| Switzerland | 221 | 203 | 424 | 1 | 42 | 43 |
| Turkey | -- | -- | -- | 95 | 356 | 451 |
| United Kingdom | 5348 | 1814 | 7162 | 386 | 629 | 1015 |
| United States | 3855 | 1423 | 5278 | 72 | 238 | 310 |
| Total | 23,503 | 12,587 | 36,090 | 1974 | 3641 | 5615 |

The 27 strata were independently imputed, allowing for the exploration of genetic variations within each stratum. After imputation, imputed variants with a Minor Allele Frequency (MAF) less than 1% were excluded. Additionally, SNPs with an INFO score below 0.7 were also excluded (Supplementary Table 2). Further details about quality controls and imputation are available in the supplementary material. To investigate the association of the resulting SNPs with ALS risk, genome-wide logistic regression analyses were conducted separately for each stratum.

### Independent test set

An external test set comprising 5615 samples (3641 cases and 1974 controls, with 3244 male and 2371 female participants) from 9 countries (Table 1) was utilised for an independent evaluation of the model’s performance. These samples, collected between 2016 and 2021, underwent genotyping, imputation and quality control by Project MinE in 2021. Quality control and imputation procedures were described in the original article [15], and are briefly summarised in the supplementary material.

### Data reliability test: GWAS and SNP-based heritability estimates

To ensure the reliability of the imputed training data, we repeated the ALS GWAS and compared the results with those published [14]. We applied logistic regression under an additive model to each stratum. To account for potential confounding factors and population structure, the first ten principal components were introduced as covariates. Individual strata underwent subsequent integration through an inverse-variance-weighted, fixed-effect meta-analysis. Moreover, we calculated the genomic inflation factor, a measure of test statistic inflation due to population stratification or other sources, and generated quantile-quantile and Manhattan plots. Finally, the SNP-based heritability was estimated. Genetic relationship matrices (GRMs) were generated incorporating genotype dosages passing quality control in all strata. Related pairs with a relatedness threshold (off-diagonal) exceeding 0.05 were excluded based on the diagonal of the Genetic relationship matrix (GRM). The first ten principal components’ eigenvectors were included as fixed effects to address population structure. The prevalence of ALS was defined as the lifetime risk for ALS (1 in 300) [4]. Estimation of SNP-based heritability for non-genome-wide-significant SNPs involved modelling genotypes for significant SNPs as fixed effects, allowing the GRM variance to reflect the heritability of all non-genome-wide-significant SNPs. We used PLINK v1.9 [35] and Genome-wide Complex Trait Analysis software (GCTA) v1.94.1 [36] for these analyses.

### Majority imputation of missing genotypes

As expected, some SNPs were missing across strata given that nine different genotyping arrays were used. Because of the potential for bias introduced by the patterns of missing values, particularly in predictive modelling where missingness might inadvertently be interpreted as a feature, a majority imputation technique was used. We replaced missing values in all datasets with the most frequently occurring value at the respective position across the training set. This approach ensures that missing values are filled based on the pattern within the whole training data, thereby minimizing the risk of introducing bias. By employing majority imputation, we aimed to preserve the integrity of the dataset and mitigate the potential impact of missingness on subsequent analyses.

### Dimensionality reduction with Gene-PCA

The abundance of features, in this case SNPs, within a dataset poses significant challenges for potential overfitting in both machine learning and deep learning methodologies [32]. To address this challenge, we used a dimensionality reduction approach. In adherence to the DiseaseCapsule protocol, we selected SNPs with a p-value less than 0.05, as determined by referencing the original GWAS [34]. Following the identification of relevant SNPs, the ANNOVAR tool [37] was used to perform SNP annotation and assign SNPs to genes through ‘gene-based annotation’ using the human reference genome hg19, and the NCBI Reference Sequence (RefSeq) database [38]. Principal Component Analysis (PCA) for each gene (Gene-PCA) was used to further streamline the feature set. The number of principal components per gene ‘k’ in PCA was adjusted according to ‘n’ the number of available SNPs per gene, tailoring the analysis to the unique characteristics of the dataset: *k*⍰= ⍰8 for *n*⍰> ⍰20, *k*⍰= ⍰4 for 4⍰< ⍰*n*⍰≤ ⍰20 and *k*⍰= ⍰1 for 1⍰≤ ⍰*n*⍰≤ ⍰4. This parameter adjustment ensured that PCA was applied with varying degrees of dimensionality reduction based on the size of the input dimension ‘n’. The PCA process was deliberately performed on the training set data alone and the learned PCA transformation was then applied to the validation and test set data deviating from the original protocol. This approach secures the repeatability of the analysis on external samples and ensures that the dimensionality reduction process remains agnostic of the validation and test sets.

### Modelling

DiseaseCapsule [32] was used as neural network architecture to build a classifier with binary-valued output for cases and controls. Resampling was employed to augment the training datasets, achieving a balanced 1:1 ratio of cases to controls. Validation involved leave-one-out and leave-half-out methods to evaluate model performance on a balanced dataset (details below). All hyperparameters were fine-tuned to enhance its training dynamics (see supplementary materials).

### Leave-one-out method

The leave-one-out method employed in this study involves the construction of twelve models. There are twelve different national strata in total from Project MinE (the Portuguese dataset was removed because of its small sample size). In each model, eleven strata served as the training set, and the remaining one was utilized as the validation set, effectively leaving all data from one country out from the training. This process was repeated until each country had been used as the validation set. This approach allows for a comprehensive evaluation of the model’s performance, ensuring that each data point has the opportunity to serve as both training and validation instances, and that eventual population effects are evaluated.

### Leave-half-out method

The leave-half-out method adopted in this study involves the construction of twelve models, where, in each model, eleven strata plus half of the remaining dataset were utilized as the training set, and the other half stratum served as the validation set. The leave-half-out approach ensures that the model is exposed to all diverse populations during training, providing a thorough assessment of its performance across various training and testing scenarios accounting for population differences.

### Population screening evaluation

To assess the performance of our model in identifying people at risk of developing ALS in the general population, we adopt a ratio of 1:300 for cases to controls, mirroring the lifetime prevalence rates observed in the population [4]. 30 cases and 9000 controls were selected for the validation set, while the remaining data was utilized for model training. We conducted 10 repetitions using randomly selected cases and controls each time.

### Evaluation metrics

We used the following standard measures to evaluate the performance of the classifiers: accuracy, precision, recall, F1 score, and the area under the receiver operating characteristic curve (AUC). Their definitions are available in the supplementary methods.

## Results

### Data reliability test

In our GWAS, SNPs in four loci were significantly associated with ALS after Bonferroni genome-wide correction (p-value < 5 × 10^−8^) (Figure 1b), which is consistent with the results published in 2016 (Figure 1a) using the same genotyping data [39]. Reasonable inflation of test statistics in the quantile–quantile plot was observed in our study (λ_GC_ = 1.015) (Supplementary Figure 1). We estimated the SNP-based heritability at 9.2%, which is in line with that published ranging from 7.2% to 9.5% [34].

**Figure 1.**
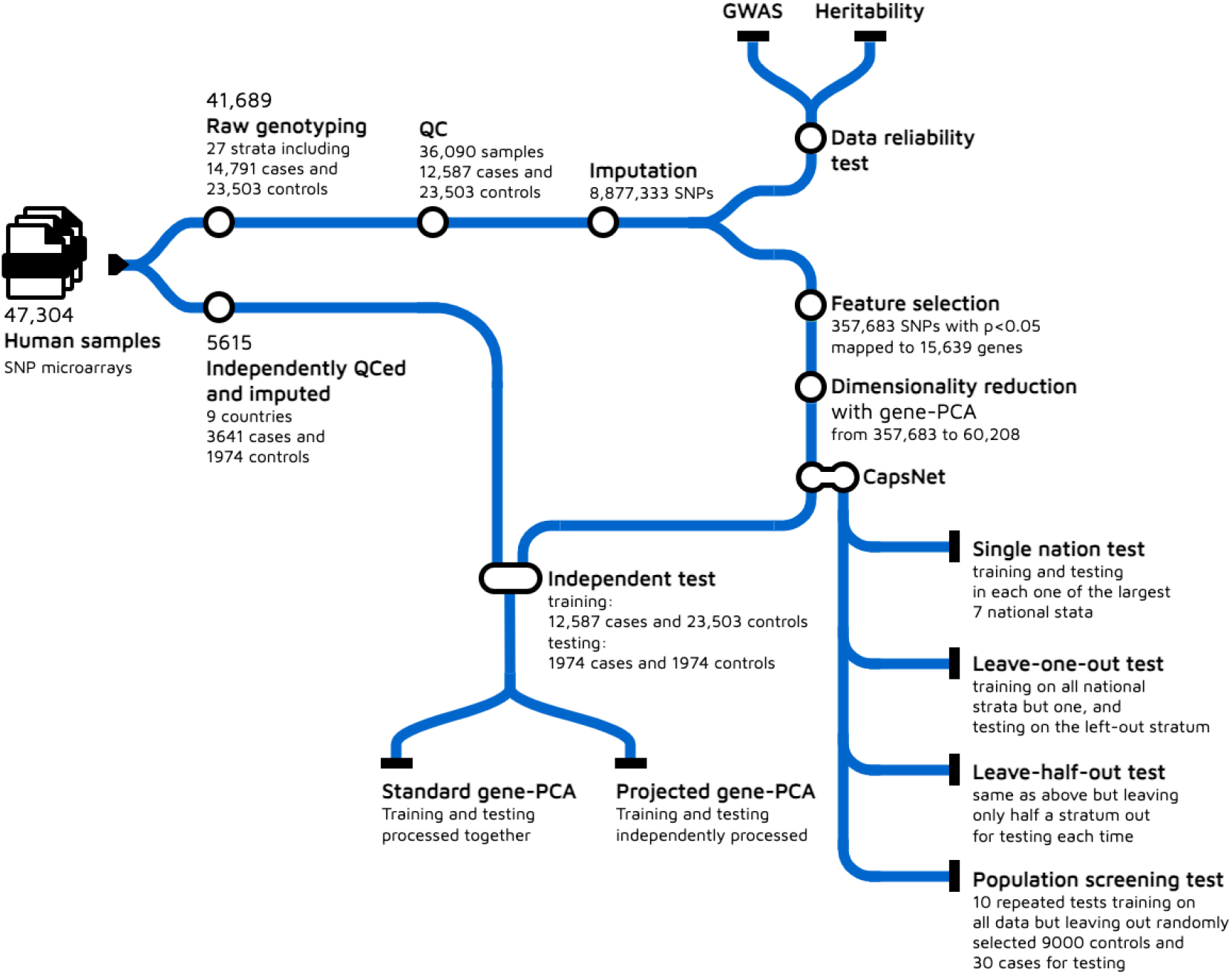
An overview of the study workflow.

**Figure 1a.**
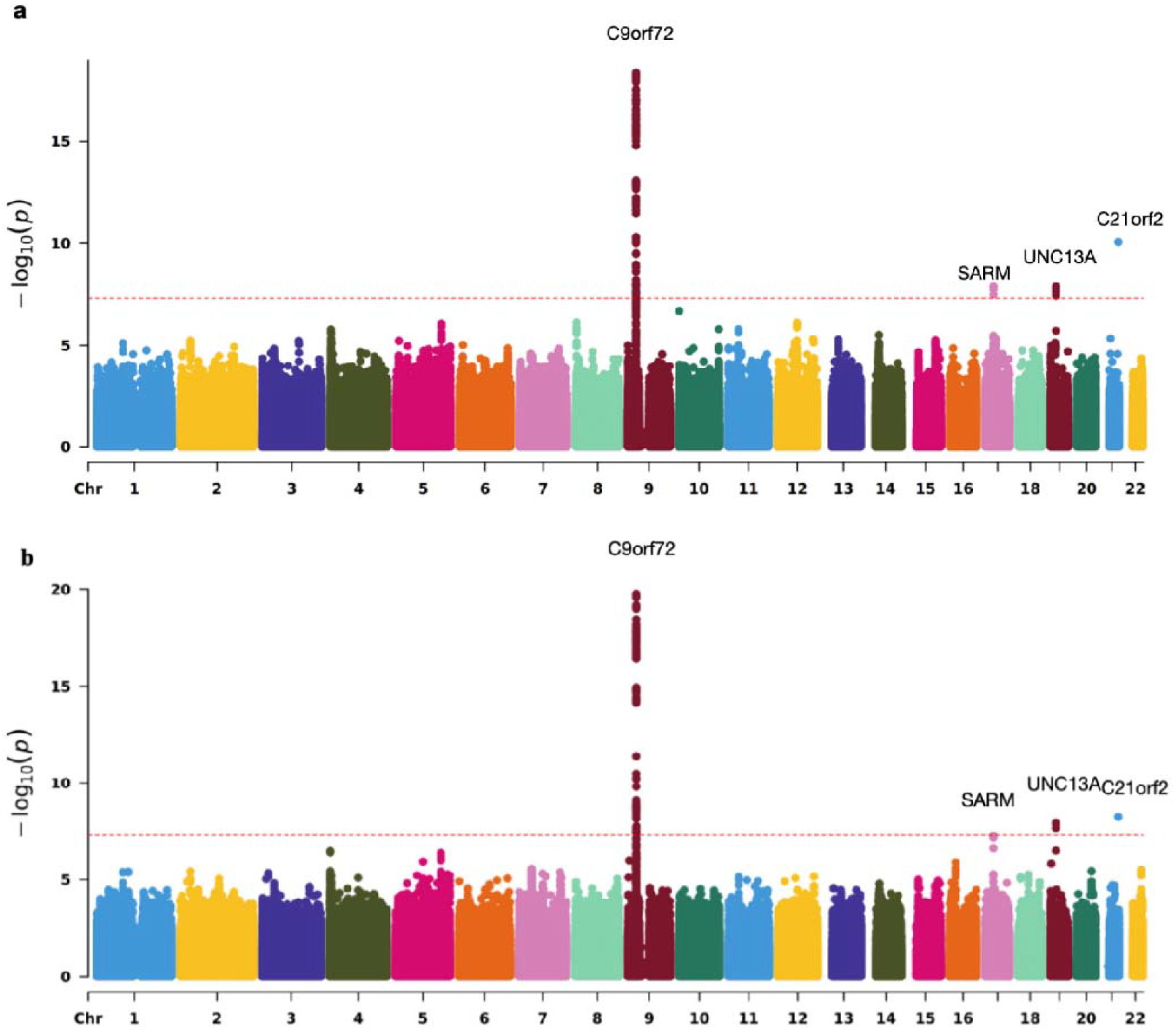
Manhattan plot for the meta-analysis results a) of the 2016 published GWAS paper, and of b) our study.

### Feature selection

There were 8,877,333 SNPs in the imputed data after merging 27 strata altogether. Following the original protocol, the 451,909 SNPs with p-value < 0.05 were selected. Using ANNOVAR 357,683 significant SNPs were mapped to 15,639 genes. Gene-PCA for all genes produced 60,208 principal components.

### Performance of single strata

The initial evaluation focused on assessing the model’s performance within individual national cohorts for which at least 1200 samples were available, with a comparison on its performance in the Dutch dataset used in the original study [32]. The results of this analysis, detailed in Table 2, underscore the model’s success in our Dutch stratum (F1 = 0.816 and AUC = 0.789). However, the performance in the other national cohorts was generally suboptimal. As the Dutch dataset was both the largest and the one with the best performance, we assessed whether this could be explained by considering sample size alone across the national datasets. Excluding the Dutch cohort, we performed Pearson’s correlation tests between the total number of samples and all performance metrics across the national cohorts and found no significant correlations (all p-values > 0.05).

**Table 2.** Performance metrics for single national stratum evaluation.

| Stratum | Train Controls | Train Cases | Validation Controls | Validation Cases | Total samples | Accuracy | Precision | Recall | F1 | AUC |
| --- | --- | --- | --- | --- | --- | --- | --- | --- | --- | --- |
| The Netherlands | 7191 | 1627 | 489 | 489 | 9796 | 0.785 | 0.714 | 0.953 | 0.816 | 0.789 |
| United Kingdom | 4991 | 1457 | 357 | 357 | 7162 | 0.566 | 0.559 | 0.627 | 0.591 | 0.560 |
| United States | 3593 | 1161 | 262 | 262 | 5278 | 0.542 | 0.548 | 0.481 | 0.512 | 0.518 |
| France | 1552 | 375 | 107 | 107 | 2141 | 0.505 | 1.000 | 0.009 | 0.018 | 0.549 |
| Germany | 766 | 1776 | 141 | 141 | 2824 | 0.564 | 0.534 | 1.000 | 0.696 | 0.561 |
| Ireland | 707 | 505 | 67 | 67 | 1346 | 0.567 | 0.569 | 0.552 | 0.560 | 0.563 |
| Italy | 1222 | 2198 | 190 | 190 | 3800 | 0.534 | 0.523 | 0.774 | 0.624 | 0.541 |
| Luo et al. (NL) | 4635 | 2222 | 520 | 520 | 7897 | 0.869 | 0.852 | 0.894 | 0.872 | --- |

### Performance of leave-one-out method and leave-half-out method

To have better performance and build a model that can be used among European populations, we attempted a leave-one-out and a leave-half-out experiments. These resulted in larger training sets (Supplementary Table 3) that were more representative of the heterogenous landscape of ALS. The tests of both approaches resulted in a generally higher performance across the national cohorts (Table 3). However, while in the leave-one-out test the model showed a suboptimal performance in the British cohort, in the leave-half-out test the model’s performance was consistently high in all cohorts.

**Table 3.** Performance of leave-one-out method and leave-half-out tests.

| Name | leave-one-out |  |  |  |  | leave-half-out |  |  |  |  |
| --- | --- | --- | --- | --- | --- | --- | --- | --- | --- | --- |
|  | Accuracy | Precision | Recall | F1 | AUC | Accuracy | Precision | Recall | F1 | AUC |
| sBE | 0.823 | 0.908 | 0.720 | 0.803 | 0.905 | 0.859 | 0.910 | 0.798 | 0.850 | 0.920 |
| sCZ | 0.835 | 0.837 | 0.833 | 0.835 | 0.907 | 0.886 | 0.890 | 0.881 | 0.885 | 0.924 |
| sFIN | 0.852 | 0.924 | 0.766 | 0.838 | 0.919 | 0.867 | 0.872 | 0.861 | 0.866 | 0.945 |
| sFR | 0.812 | 0.754 | 0.927 | 0.832 | 0.894 | 0.788 | 0.926 | 0.627 | 0.748 | 0.889 |
| sGER | 0.714 | 0.848 | 0.523 | 0.647 | 0.808 | 0.687 | 0.625 | 0.932 | 0.748 | 0.792 |
| sIB | 0.828 | 0.865 | 0.778 | 0.819 | 0.855 | 0.796 | 0.809 | 0.776 | 0.792 | 0.830 |
| sIR | 0.791 | 0.956 | 0.610 | 0.745 | 0.888 | 0.830 | 0.916 | 0.727 | 0.811 | 0.924 |
| sIT | 0.831 | 0.909 | 0.736 | 0.813 | 0.911 | 0.826 | 0.776 | 0.915 | 0.840 | 0.916 |
| sNL | 0.776 | 0.845 | 0.674 | 0.750 | 0.865 | 0.804 | 0.964 | 0.632 | 0.763 | 0.918 |
| sSW | 0.776 | 0.894 | 0.626 | 0.736 | 0.858 | 0.788 | 0.863 | 0.685 | 0.764 | 0.858 |
| sUK | 0.627 | 0.782 | 0.353 | 0.486 | 0.822 | 0.810 | 0.828 | 0.784 | 0.805 | 0.877 |
| sUS | 0.801 | 0.830 | 0.758 | 0.792 | 0.886 | 0.820 | 0.880 | 0.741 | 0.805 | 0.908 |

### Results of a simulated population screening test

A ratio of 1:300 for cases to controls was maintained in the validation set, with 30 cases and 9,000 controls selected. Additionally, to enhance the reliability of our findings, 10 repetitions were conducted. This approach ensures that our results reflect real-world lifetime risk estimates and provide insights into the model’s performance in a context of population screening.

Table 4 reports the results of our analysis. Precision and F1 score were low but still acceptable given the highly unbalanced nature of the validation set. Indeed, interpreting precision as the probability of developing ALS given a positive test, this was over 7 times higher (0.024) than the probability of developing ALS in the general population (1/300 = 0.0033). This probability aligns closely with those previously estimated for people testing positive for the main ALS genes in the context of population screening when their non-complete penetrance, disease and mutation frequencies, and of the test sensitivity and specificity were taken into account (*SOD1: 0.109; FUS: 0.0302; C9orf72: 0.0198*) [40, 41].

**Table 4.** Performance of models in a simulated real population screening.

|  | Controls | Cases | Accuracy | TN/9000 | TP/30 | Precision | Recall | F1 | AUC | Notes |
| --- | --- | --- | --- | --- | --- | --- | --- | --- | --- | --- |
| 1 | 9000 | 30 | 0.892 | 0.893 | 0.767 | 0.023 | 0.767 | 0.045 | 0.830 | TP=23, TN=8033, FP=967, FN=7 |
| 2 | 9000 | 30 | 0.889 | 0.889 | 0.933 | 0.027 | 0.933 | 0.052 | 0.911 | TP=28, TN=8002, FP=998, FN=2 |
| 3 | 9000 | 30 | 0.890 | 0.890 | 0.900 | 0.027 | 0.900 | 0.052 | 0.895 | TP=27, TN=8009, FP=991, FN=3 |
| 4 | 9000 | 30 | 0.891 | 0.892 | 0.867 | 0.026 | 0.867 | 0.050 | 0.879 | TP=26, TN=8024, FP=976, FN=4 |
| 5 | 9000 | 30 | 0.885 | 0.885 | 0.733 | 0.021 | 0.733 | 0.041 | 0.809 | TP=22, TN=7969, FP=1031, FN=8 |
| 6 | 9000 | 30 | 0.887 | 0.887 | 0.867 | 0.025 | 0.867 | 0.049 | 0.877 | TP=26, TN=7984, FP=1016, FN=4 |
| 7 | 9000 | 30 | 0.888 | 0.888 | 0.800 | 0.023 | 0.800 | 0.045 | 0.844 | TP=24, TN=7995, FP=1005, FN=6 |
| 8 | 9000 | 30 | 0.882 | 0.882 | 0.767 | 0.021 | 0.767 | 0.041 | 0.824 | TP=23, TN=7938, FP=1062, FN=7 |
| 9 | 9000 | 30 | 0.890 | 0.890 | 0.700 | 0.021 | 0.700 | 0.041 | 0.795 | TP=21, TN=8012, FP=988, FN=9 |
| 10 | 9000 | 30 | 0.884 | 0.885 | 0.867 | 0.024 | 0.867 | 0.047 | 0.876 | TP=26, TN=7961, FP=1039, FN=4 |
| Mean | 9000 | 30 | 0.888 | 0.888 | 0.820 | 0.024 | 0.820 | 0.046 | 0.854 | --- |

### Performance on the independent test set

An evaluation was conducted on an external dataset to assess the model’s performance on independent data. In this experiment the full dataset from the 2016 ALS study (12,587 cases and 23,503 controls) was used for training, while balanced subsets of the whole independent dataset (1,976 cases and 1,976 controls) and of its individual national cohorts for which at least 100 cases and controls were available, were used for testing. Table 5 provides details on the numbers of cases and controls within each cohort, along with the corresponding performance metrics. Figure 3 illustrates the Receiver Operating Characteristic (ROC) curves for the various cohorts. In this evaluation, two distinct approaches to Gene-PCA were explored. The first approach involved applying Gene-PCA to both the training and test datasets simultaneously, as in the original implementation of the method. However, this method carries a risk of overfitting, as it may cause the principal components (PCs) to explain more variance in the test set than they typically would in a truly independent dataset. Moreover, it makes analysing new external samples impossible without re-processing the whole data from scratch and re-train the model. To overcome these factors a second approach was adopted. In this alternative method, Gene-PCA was performed exclusively on the training set. The PCs derived from the training set were then projected onto the test set, ensuring that the test data remained completely unseen during the model’s development. This separation helps preserve the integrity and generalizability of the model. No substant performance differences were observed between the two approaches. Overall, the performance was satisfactory across the five national strata (0.64 < AUC < 0.90), with an AUC of 0.79 for the entire dataset.

**Table 5.**
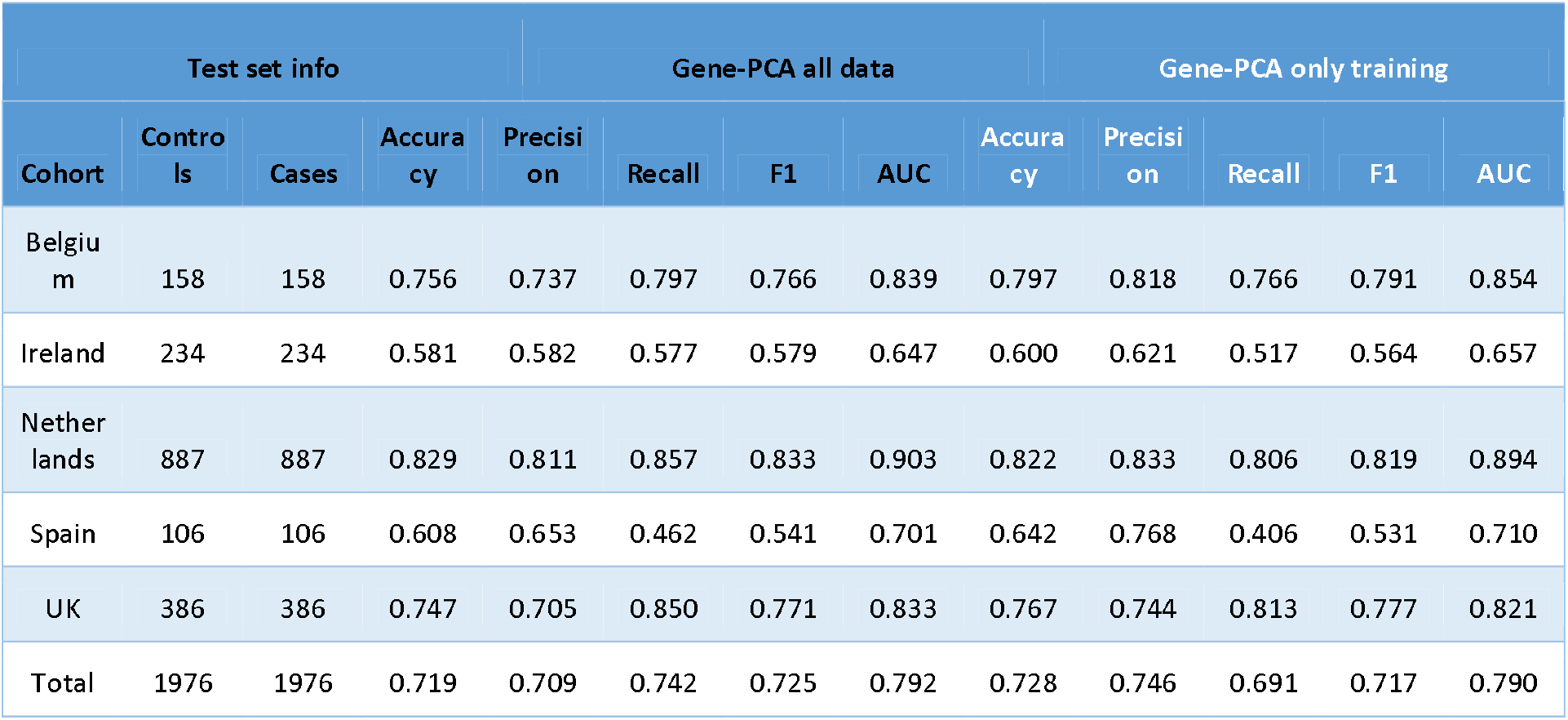
Number of cases and controls from different cohorts and their performance.

**Figure 3.**
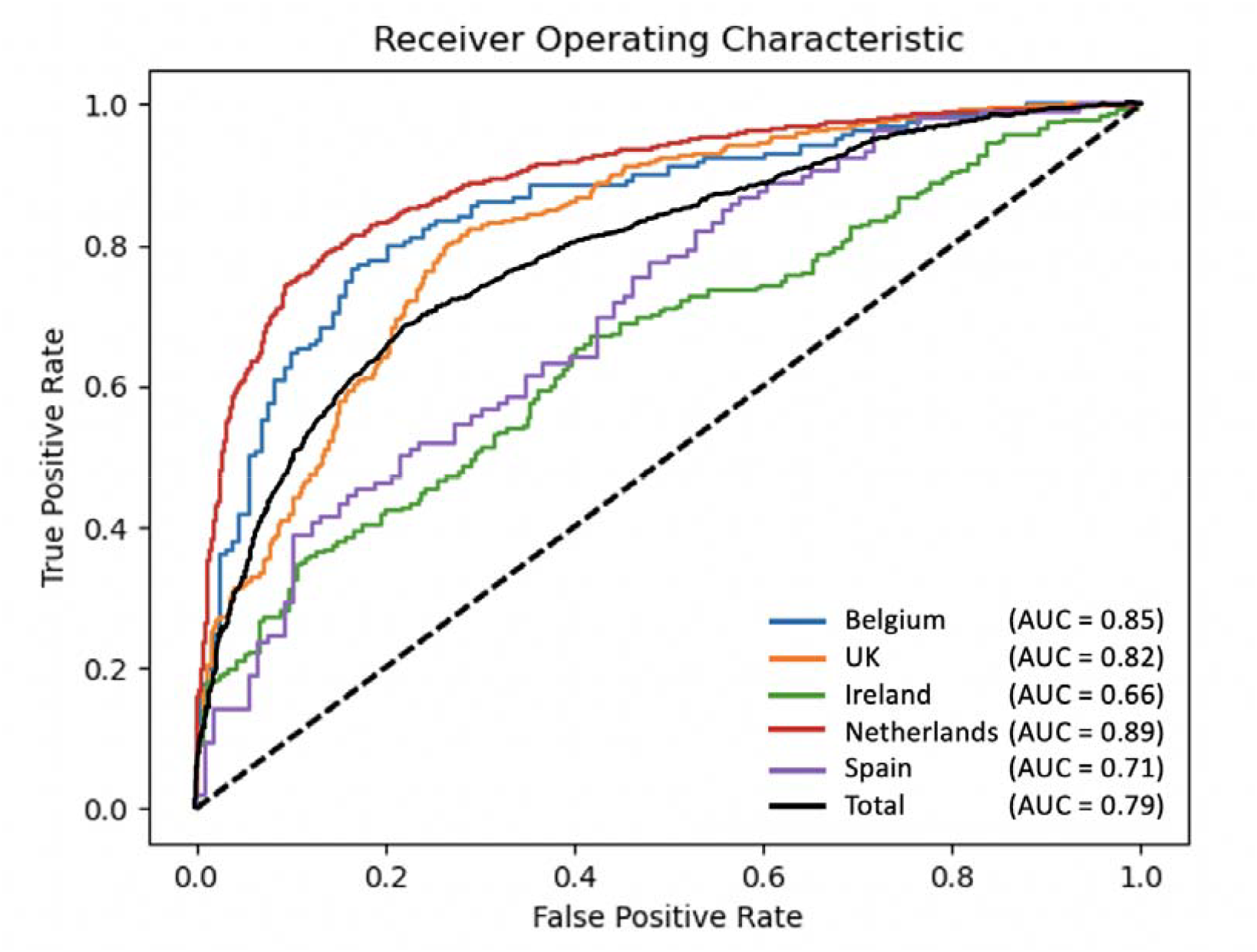
ROC and AUC of different cohorts and the total dataset with gene-PCA performed only on the training data.

### Performance of other methods

To comprehensively evaluate the performance of our model in comparison to other methods, we tested several diverse modelling techniques (Table 6) using the same training and testing datasets as in the previous experiment. The PRS-based models were constructed by selecting SNPs using publicly available GWAS summary statistics from the 2016 set [14] for consistency with the training set used for the machine learning methods and to ensure the independency of the test set, with significance thresholds of P⍰< ⍰5l1×⍰10−2, P⍰< ⍰5⍰× ⍰10−4, P⍰< ⍰5⍰× ⍰10−6, and P⍰< ⍰5⍰× ⍰10−8, respectively. Additionally, logistic regression (LR), random forest (RF), and convolutional neural network (CNN) methods were applied, each utilizing the same feature sets and sample cohorts as the CapsNets. The accuracy, F1 and AUC for CapsNet were higher than for any other methods (Table 6).

**Table 6.** Classification results for external test set. LR, logistic regression; RF, Random Forest; CNN, Convolutional Neural Network. a, b, c and d represent PRS-based models that the SNPs were selected by GWAS with the threshold P⍰< ⍰5⍰× ⍰10^−8^, P⍰< ⍰5⍰× ⍰10^−6^, P⍰< ⍰5⍰× ⍰10^−4^ and P⍰< ⍰5⍰× ⍰10 ^−2^, respectively.

| Method | Accuracy | Precision | Recall | F1 | AUC |
| --- | --- | --- | --- | --- | --- |
| PRS<sup>a</sup> | 0.520 | 0.551 | 0.222 | 0.316 | 0.531 |
| PRS<sup>b</sup> | 0.497 | 0.499 | 0.988 | 0.663 | 0.524 |
| PRS<sup>c</sup> | 0.554 | 0.664 | 0.218 | 0.328 | 0.626 |
| PRS<sup>d</sup> | 0.676 | 0.782 | 0.487 | 0.600 | 0.785 |
| LR | 0.524 | 0.513 | 0.994 | 0.677 | 0.784 |
| RF | 0.521 | 0.518 | 0.619 | 0.564 | 0.538 |
| CNN | 0.582 | 0.600 | 0.489 | 0.539 | 0.629 |
| CapsNet | 0.728 | 0.746 | 0.691 | 0.717 | 0.790 |

## Discussion

In this study, we conducted a comprehensive investigation into the generalizability and performance of a promising deep-learning method for predicting which people have an increased risk of ALS based on their SNP genotypes as assayed by microarrays. The resulting model overcomes the limits of a previous, non-generalizable implementation by training and testing on large international datasets that include thousands of people with ALS and controls from 11 European countries and the US. Furthermore, the method can now be applied in real research and clinical contexts, as it can be used on external samples not pre-processed with the training dataset, and can accommodate genotyping arrays from various technologies, including those with missing genotypes.

In our first experiment (Table 1), we evaluated the model’s performance using seven cohorts from different European countries and the US, training and validating on each cohort separately. The model demonstrated optimal performance in the Dutch cohort consistent with the original study [32], as evident from the high accuracy, precision, recall, F1 score, and AUC values. However, the transferability of the model to other cohorts appeared limited with suboptimal performance in all other national datasets (AUC < 0.6). This experiment emphasized the importance of further investigation and potential adaptations for enhanced generalizability.

The better performance of the initial Dutch model in the original study was possibly related to its size as this dataset had a larger number of cases and controls (6857 individuals in total) than the other national cohorts in our study, providing the model with a larger dataset for learning that would have contributed to their model performance. However, as the model’s performance metrics in the other cohorts did not correlate with their size and the model performed poorly on datasets of similar sizes such as the UK and US datasets (6448 and 4754 individuals respectively), a role might have been played by the specific population structure of the Dutch cohort [42], which could translate in a limited applicability to other populations.

To address this, we next evaluated models trained on an international larger and more diverse dataset comprising tens of thousands of cases and controls from 12 countries (Table 3). The models demonstrated significantly improved performance (0.78 < AUC < 0.96) both when validated on the national cohorts excluded from training (leave-one-out) and when all national cohorts were included in the training set (leave-half-out). The performance of the models improved consistently across all cohorts when all populations were included in the training set (leave-half-out) with a notable increase in recall for several populations, underscoring the importance of a diverse, multinational training set for maximizing model consistency and generalizability.

In the subsequent experiment, we tested the model’s performance in a scenario mimicking population screening, where the lifetime risk of ALS is 1 in 300, resulting in highly imbalanced validation datasets (9000 controls and 30 cases; Table 4). Despite this imbalance, the models demonstrated high performance metrics (0.80 < AUC < 0.91). However, the average precision was very low (0.024), as expected given the stark imbalance between cases and controls. This precision rate aligns closely with estimates for individuals testing positive for the major ALS genes in population screening (*SOD1: 0*.*109; FUS: 0*.*0302; C9orf72: 0*.*0198) [*40]. When interpreting this result, it is important to consider that such low probabilities are due to four key factors: the incomplete penetrance of most ALS genetic variants (SOD1: 0.70; FUS: 0.58; C9orf72: 0.44), the rarity of ALS, the small proportion of patients who carry these variants (SOD1: 0.02; FUS: 0.004; C9orf72: 0.06), and the performance of the genetic test (sensitivity and specificity)[40, 41, 43]. It is also important to consider that a direct consequence of the imbalance is that the number of individuals who might be at high risk of developing ALS within the true negative set (the controls) may be similar to the number of true positives (the cases) in the validation set. This could lead to an overestimation of the false positives and an underestimation of the model performance in our experiment.

To further evaluate the robustness of our method, we tested a model trained on a large international dataset (over 40,000 individuals from 12 countries) on an independent international dataset of approximately 4,000 cases and controls from nine European populations and the US. This independent dataset was imputed and underwent quality control following different protocols due to its origin in a separate study, representing a more challenging testing scenario. Overall, the performance was satisfactory across the five larger national strata (0.66 < AUC < 0.89), and in the entire dataset with an AUC of 0.79.

Finally, we compared our model with other machine learning methods and polygenic risk score (PRS) approaches using the same independent dataset. Our model consistently outperformed the other methods, showing a particularly substantial improvement (∼40%) in recall compared to PRS. This improvement may be due to the ability of deep learning to capture complex interactions and non-linear effects across genetic variants translating into a larger proportion of people with ALS identified by the model.

In the era of precision medicine, DNA sequencing plays a crucial role in supporting diagnostics, tailoring healthcare, and identifying individuals at risk of developing diseases. In ALS, traditional approaches based on genotyping disease-causing variants in known disease genes are currently applicable or effective only for a minority of people with ALS. These include individuals with a family history of the disease or the 10–20% of sporadic cases carrying a known disease-causing mutation. PRSs could, in principle, be applied to all individuals; however, they have shown limited translational success in ALS due to their restricted ability to capture disease risk and identify individuals at risk. In this context, our approach could address a significant part of this shortfall by supporting effective diagnostic strategies and screening programs for all individuals at risk.

Despite the potential of our method, some limitations remain, particularly the restriction of model evaluation to European populations, excluding individuals of Asian, Central and South American, or African descent. Unfortunately, large datasets from these populations were unavailable to us during this study. To address this, we are collaborating with international ALS consortia, such as Project MinE, to collect more diverse samples from underrepresented populations, which will help refine and expand the applicability of our method. Another factor to consider when comparing our approach to classic genetic testing is the lack of gene-specific information it provides, which limits its use for guiding gene therapy strategies.

In conclusion, this study demonstrates the potential of deep learning methods for developing predictive models of ALS. While further development is needed, our method lays the foundation for a genetic test with diagnostic potential that could be applied to all individuals regardless of their family history or the presence of known ALS mutations.

## Data availability

All data used in this study are publicly available. The ALS genome data and GWAS data used in this study are from Project MinE and can be accessed via online application (www.projectmine.com). Other data utilized in this study include the following: the Wellcome Trust Case Control Consortium (https://www.wtccc.org.uk/) and dbGaP datasets (phs000101.v3.p1, phs000101.v3.p1, phs000101.v3.p1, phs000101.v3.p1, phs000126.v1.p1, phs000196.v1.p1, phs000344.v1.p1, phs000344.v1.p1, phs000344.v1.p1).

## Acknowledgements

Samples used in this research were in part obtained from the UK National DNA Bank for MND Research, funded by the MND Association and the Wellcome Trust. Part of the samples were obtained from The Project MinE and MND centres internationally. We thank people with MND and their families for their participation in this project. The authors acknowledge use of the King’s Computational Research, Engineering and Technology Environment (CREATE) (https://create.kcl.ac.uk), which is delivered in partnership with the National Institute for Health and Care Research (NIHR) Biomedical Research Centres at South London and Maudsley and Guy’s and St. Thomas’ NHS Foundation Trusts and part-funded by capital equipment grants from the Maudsley Charity (award 980) and Guy’s and St. Thomas’ Charity (TR130505). We also acknowledge Health Data Research UK, which is funded by the UK Medical Research Council, Engineering and Physical Sciences Research Council, Economic and Social Research Council, Department of Health and Social Care (UK), Chief Scientist Office of the Scottish Government Health and Social Care Directorates, Health and Social Care Research and Development Division (Welsh Government), Public Health Agency (Northern Ireland), British Heart Foundation and Wellcome Trust. The authors would like to thank Mr Renato Santos from King’s College London for its help with the article’s figures. Several authors of this publication are member of the European Reference Network for Neuromuscular Diseases – Project ID N° 870177.

## Reference

1. Brown, R.H. and A. Al-Chalabi, Amyotrophic lateral sclerosis. New England Journal of Medicine, 2017. 377(2): p. 162–172.

2. Mata, S., et al., Epidemiology of amyotrophic lateral sclerosis in the north east Tuscany in the 2018–2021 period. Eneurologicalsci, 2023. 31: p. 100457.

3. Longinetti, E. and F. Fang, Epidemiology of amyotrophic lateral sclerosis: an update of recent literature. Current opinion in neurology, 2019. 32(5): p. 771–776.

4. Johnston, C.A., et al., Amyotrophic lateral sclerosis in an urban setting. Journal of neurology, 2006. 253(12): p. 1642–1643.

5. Al-Chalabi, A., et al., An estimate of amyotrophic lateral sclerosis heritability using twin data. Journal of Neurology, Neurosurgery & Psychiatry, 2010. 81(12): p. 1324–1326.

6. Shatunov, A. and A. Al-Chalabi, The genetic architecture of ALS. Neurobiology of disease, 2021: p. 105156.

7. Mehta, P.R., et al., The impact of age on genetic testing decisions in amyotrophic lateral sclerosis. Brain, 2022. 145(12): p. 4440–4447.

8. Dilliott, A.A., et al., Clinical testing panels for ALS: global distribution, consistency, and challenges. Amyotrophic lateral sclerosis and frontotemporal degeneration, 2023: p. 1–16.

9. Abel, O., et al., ALSoD: A user-friendly online bioinformatics tool for amyotrophic lateral sclerosis genetics. Hum Mutat, 2012. 33(9): p. 1345–51.

10. Iacoangeli, A., et al., Oligogenic structure of amyotrophic lateral sclerosis has genetic testing, counselling and therapeutic implications. Journal of Neurology, Neurosurgery & Psychiatry, 2025.

11. Iacoangeli, A., et al., ALSgeneScanner: a pipeline for the analysis and interpretation of DNA sequencing data of ALS patients. Amyotroph Lateral Scler Frontotemporal Degener, 2019. 20(3-4): p. 207–215.

12. Renton, A.E., A. Chiò, and B.J. Traynor, State of play in amyotrophic lateral sclerosis genetics. Nat Neurosci, 2014. 17(1): p. 17–23.

13. Iacoangeli, A., et al., Genome-wide Meta-analysis finds the ACSL5-ZDHHC6 locus Is associated with ALS and links weight loss to the disease genetics. Cell reports, 2020. 33(4): p. 108323.

14. van Rheenen, W., et al., Genome-wide association analyses identify new risk variants and the genetic architecture of amyotrophic lateral sclerosis. Nature Genetics, 2016. 48(9): p. 1043–1048.

15. van Rheenen, W., et al., Common and rare variant association analyses in amyotrophic lateral sclerosis identify 15 risk loci with distinct genetic architectures and neuron-specific biology. Nature Genetics, 2021. 53(12): p. 1636–1648.

16. Nicolas, A., et al., Genome-wide analyses identify KIF5A as a novel ALS gene. Neuron, 2018. 97(6): p. 1268–1283. e6.

17. Restuadi, R., et al., Polygenic risk score analysis for amyotrophic lateral sclerosis leveraging cognitive performance, educational attainment and schizophrenia. European Journal of Human Genetics, 2022. 30(5): p. 532–539.

18. Pain, O., et al., Harnessing transcriptomic signals for amyotrophic lateral sclerosis to identify novel drugs and enhance risk prediction. Heliyon, 2024. 10(15).

19. Balvert, M., et al., Considerations in the search for epistasis. Genome Biology, 2024. 25(1): p. 296.

20. Bean, D.M., et al., A Knowledge-Based Machine Learning Approach to Gene Prioritisation in Amyotrophic Lateral Sclerosis. Genes (Basel), 2020. 11(6).

21. Grollemund, V., et al., Manifold learning for amyotrophic lateral sclerosis functional loss assessment. Journal of Neurology, 2021. 268(3): p. 825–850.

22. Hu, J., et al., DGLinker: flexible knowledge-graph prediction of disease–gene associations. Nucleic Acids Research, 2021. 49(W1): p. W153–W161.

23. Marriott, H., et al., Unsupervised machine learning identifies distinct ALS molecular subtypes in post-mortem motor cortex and blood expression data. Acta Neuropathol Commun, 2023. 11(1): p. 208.

24. Tavazzi, E., et al., Predicting functional impairment trajectories in amyotrophic lateral sclerosis: a probabilistic, multifactorial model of disease progression. Journal of Neurology, 2022. 269(7): p. 3858–3878.

25. Zhang, S., et al., Genome-wide identification of the genetic basis of amyotrophic lateral sclerosis. Neuron, 2022. 110(6): p. 992–1008.e11.

26. O’Neill, K., et al., ALS molecular subtypes are a combination of cellular and pathological features learned by deep multiomics classifiers. Cell Reports, 2025. 44(3).

27. Spargo, T.P., et al., Unsupervised machine-learning identifies clinically distinct subtypes of ALS that reflect different genetic architectures and biological mechanisms. medRxiv, 2023: p. 2023.06. 12.23291304.

28. Müller, M., et al., Explainable models of disease progression in ALS: Learning from longitudinal clinical data with recurrent neural networks and deep model explanation. Computer Methods and Programs in Biomedicine Update, 2021. 1: p. 100018.

29. van der Burgh, H.K., et al., Deep learning predictions of survival based on MRI in amyotrophic lateral sclerosis. NeuroImage: Clinical, 2017. 13: p. 361–369.

30. Yin, B., et al., Using the structure of genome data in the design of deep neural networks for predicting amyotrophic lateral sclerosis from genotype. Bioinformatics, 2019. 35(14): p. i538–i547.

31. Sabour, S., N. Frosst, and G.E. Hinton, Dynamic routing between capsules.. 2017.

32. Luo, X., X. Kang, and A. Schönhuth, Predicting the prevalence of complex genetic diseases from individual genotype profiles using capsule networks. Nature Machine Intelligence, 2023. 5(2): p. 114–125.

33. Byrne, R.P., et al., Dutch population structure across space, time and GWAS design. Nature communications, 2020. 11(1): p. 4556.

34. van Rheenen, W., et al., Genome-wide association analyses identify new risk variants and the genetic architecture of amyotrophic lateral sclerosis. Nat Genet, 2016. 48(9): p. 1043–8.

35. Purcell, S., et al., PLINK: a tool set for whole-genome association and population-based linkage analyses. The American journal of human genetics, 2007. 81(3): p. 559–575.

36. Yang, J., et al., GCTA: a tool for genome-wide complex trait analysis. The American Journal of Human Genetics, 2011. 88(1): p. 76–82.

37. Wang, K., M. Li, and H. Hakonarson, ANNOVAR: functional annotation of genetic variants from high-throughput sequencing data. Nucleic Acids Res, 2010. 38(16): p. e164.

38. O’Leary, N.A., et al., Reference sequence (RefSeq) database at NCBI: current status, taxonomic expansion, and functional annotation. Nucleic Acids Res, 2016. 44(D1): p. D733–45.

39. Van Rheenen, W., et al., Genome-wide association analyses identify new risk variants and the genetic architecture of amyotrophic lateral sclerosis. Nature genetics, 2016. 48(9): p. 1043.

40. Spargo, T., et al., Modelling population genetic screening in rare neurodegenerative diseases. 2023.

41. Biesecker, L.G., Genomic screening and genomic diagnostic testing—two very different kettles of fish. Genome Medicine, 2019. 11: p. 1–3.

42. Byrne, R.P., et al., Dutch population structure across space, time and GWAS design. Nature Communications, 2020. 11(1): p. 4556.

43. Spargo, T.P., et al., Calculating variant penetrance from family history of disease and average family size in population-scale data. Genome Medicine, 2022. 14(1): p. 141.

