## Supplementary Materials for "Towards a diagnostic test for sporadic ALS utilising deep learning and SNP microarrays"

Supplementary Table 1. Description of the cohorts before quality control.

| Cohort | Name | Country | Cases | Controls | SNP | Platform |
| --- | --- | --- | --- | --- | --- | --- |
| 1 | NL1 | The Netherlands | 461 | 450 | 317,503 | Illumina317K |
| 2 | BE1 | Belgium | 311 | 371 | 370,404 | Illumina370K |
| 3 | NL2 | The Netherlands | 582 | 629 | 370,404 | Illumina370K |
| 4 | SW1 | Sweden | 493 | 500 | 370,404 | Illumina370K |
| 5 | NL3 | The Netherlands | 0 | 5,974 | 561,466 | Illumina550K |
| 6 | FR1 | France | 251 | 724 | 307,790 | Illumina317K |
| 7 | UK1 | United Kingdom | 245 | 221 | 307,790 | Illumina317K |
| 8 | US1 | United States | 753 | 811 | 307,790 | Illumina317K |
| 9 | IR1 | Ireland | 221 | 211 | 561,466 | Illumina550K |
| 10 | IR2 | Ireland | 103 | 127 | 620,901 | Illumina610K |
| 11 | UK2 | United Kingdom | 661 | 0 | 584,414 | Illumina550K |
| 12 | US2 | United States | 0 | 527 | 561,466 | Illumina550K |
| 13 | US3 | United States | 276 | 0 | 555,351 | Illumina550K |
| 14 | IT1 | Italy | 141 | 0 | 620,901 | Illumina610K |
| 15 | IT2 | Italy | 261 | 246 | 555,352 | Illumina550K |
| 16 | UK3 | United Kingdom | 0 | 2,501 | 934,848 | Illumina1M |
| 17 | UK4 | United Kingdom | 0 | 2,699 | 934,010 | Illumina1M |
| 18 | US4 | United States | 0 | 867 | 344,301 | Illumina370K |
| 19 | US5 | United States | 0 | 1,986 | 1,012,895 | Illumina1M |
| 20 | FIN1 | Finland | 0 | 201 | 346,831 | Illumina370K |
| 21 | FIN2 | Finland | 401 | 191 | 345,111 | Illumina370K |
| 22 | FIN3 | Finland | 0 | 103 | 1,199,187 | Illumina1M |
| 23 | IT3 | Italy | 1,792 | 1,107 | 545,914 | Illumina660W |
| 24 | FR2 | France | 0 | 1,100 | 582,892 | Illumina550K |
| 25 | GER1 | Germany | 0 | 677 | 561,466 | Illumina550K |
| 26 | NL4 | The Netherlands | 1,226 | 2,262 | 719,665 | IlluminaOmniExpress |
| 27 | GER2 | Germany | 580 | 286 | 719,665 | IlluminaOmniExpress |
| 28 | IT4 | Italy | 311 | 100 | 719,665 | IlluminaOmniExpress |
| 29 | PU1 | Portugal | 40 | 54 | 719,665 | IlluminaOmniExpress |
| 30 | SP1 | Spain | 105 | 63 | 719,665 | IlluminaOmniExpress |
| 31 | SWISS1 | Switzerland | 228 | 236 | 719,665 | IlluminaOmniExpress |
| 32 | BE2 | Belgium | 225 | 250 | 719,665 | IlluminaOmniExpress |
| 33 | FIN4 | Finland | 144 | 0 | 719,665 | IlluminaOmniExpress |
| 34 | IR3 | Ireland | 268 | 478 | 719,665 | IlluminaOmniExpress |
| 35 | SW2 | Sweden | 281 | 271 | 719,665 | IlluminaOmniExpress |
| 36 | US6 | United States | 65 | 42 | 964,193 | IlluminaOmniExpress |
| 37 | GER3 | Germany | 1,519 | 0 | 529,948 | IlluminaCoreExome |
| 38 | FR3 | France | 363 | 0 | 730,525 | IlluminaOmniExpress |
| 39 | US7 | United States | 573 | 0 | 730,525 | IlluminaOmniExpress |
| 40 | UK5 | United Kingdom | 1,214 | 14 | 730,525 | IlluminaOmniExpress |
| 41 | NL5 | The Netherlands | 697 | 619 | 2,391,739 | Illumina2.5M |
| Total |  |  | 14,791 | 26,898 |  |  |

### Quality control

Quality control (QC) was first performed per cohort to remove low-quality single nucleotide polymorphisms (SNPs) and individuals using PLINK 1.9. SNPs were annotated according to dbSNP137 and mapped to the hg19 reference genome. Subsequently, multi-allelic and AT/CG SNPs were removed as well as SNPs with a call rate < 98%, fewer than 10 minor allele observations per cohort, biased missingness as determined by haplotype and non-autosomal SNPs. Individuals with gender mismatches or an excessive number of heterozygous SNPs (F < -0.2) were removed. To check for strand inconsistencies or annotation errors, allele frequencies between each cohort were compared with those observed in the European population represented in 1000 Genomes Project.

Considering the low number of overlapping SNPs between all different platforms (n = 41,689), cohorts and the presence of cohorts with cases only or controls only, cohorts were combined based on reported nationality and genotyping platform. Quality control per stratum included removal of SNPs that deviated from Hardy-Weinberg equilibrium (p < 1 × 10^-5^ and p < 1 × 10^-9^ in controls and cases respectively) and those with biased missingness between cases and controls (p < 1 × 10^-3^). Subsequently, related and duplicate individuals across all strata (pi-hat > 0.1) were removed. Individuals were projected along the first four principal components (PC) calculated on HapMap3 individuals using EIGENSTRAT. Population outliers, defined as deviation > 10 SD from the HapMap CEU population mean on PCs 1-4 or > 4SD from its stratum mean on PC1-2, were removed. After removing population outliers, PCs were recalculated on an LD-pruned set of SNPs for each stratum and again outliers (> 5SD on PC1-4) were removed. After removing all outliers, PCs were recalculated once more. Based on scree plots for each stratum the eigenvectors for the first 1-4 PCs were included in the logistic regression. To calculate genomic inflation factors per stratum, the test statistic’s empirical quantiles were obtained applying logistic regression in an additive model.

Supplementary Table 2. Data statistics from each stratum after quality control.

| Stratum | Cohort(s) | Name | Cases | Controls | SNPs after quality control | SNPs after imputation | λ (GC) |
| --- | --- | --- | --- | --- | --- | --- | --- |
| 1 | 1 | sNL1 | 423 | 420 | 300,353 | 7,389,756 | 1.013 |
| 2 | 2 | sBE1 | 299 | 317 | 313,908 | 7,186,551 | 1.025 |
| 3 | 3+5 | sNL2 | 145 | 4886 | 276,115 | 7,297,390 | 1.016 |
| 4 | 4 | sSWE1 | 294 | 279 | 319,705 | 7,547,941 | 1.033 |
| 5 | 6 | sFR1 | 155 | 654 | 279,421 | 6,917,611 | 1.02 |
| 6 | 7 | sUK1 | 168 | 159 | 264,022 | 7,160,519 | 1.034 |
| 7 | 8+18 | sUS1 | 598 | 1339 | 266,792 | 7,191,502 | 1.004 |
| 8 | 9+10 | sIR1 | 308 | 331 | 490,017 | 7,483,795 | 1.023 |
| 9 | 11+16 | sUK2 | 614 | 2687 | 504,559 | 7,450,672 | 1.015 |
| 10 | 12+13 | sUS2 | 266 | 513 | 488,196 | 7,429,928 | 1.017 |
| 11 | 14+15 | sIT1 | 383 | 244 | 474,562 | 7,088,309 | 1.021 |
| 12 | 20+21 | sFIN1 | 381 | 378 | 291,624 | 7,726,711 | 1.03 |
| 13 | 26 | sNL3 | 952 | 1841 | 610,535 | 7,547,904 | 1.011 |
| 14 | 27 | sGER1 | 518 | 258 | 614,207 | 7,429,218 | 1.018 |
| 15 | 28 | sIT2 | 290 | 93 | 614,910 | 7,250,051 | 1.026 |
| 16 | 29+30 | sIB1 | 126 | 99 | 564,159 | 7,365,744 | 1.048 |
| 17 | 31 | sSWISS1 | 203 | 221 | 600,019 | 7,367,862 | 1.037 |
| 18 | 32 | sBE2 | 205 | 242 | 617,815 | 7,513,167 | 1.032 |
| 19 | 35 | sSW2 | 232 | 235 | 613,300 | 7,632,567 | 1.032 |
| 20 | 22+33 | sFIN2 | 135 | 97 | 437,401 | 7,852,851 | 1.068 |
| 21 | 34 | sIR2 | 264 | 443 | 620,690 | 7,482,021 | 1.013 |
| 22 | 19+36+39 | sUS3 | 559 | 2003 | 545,659 | 7,468,102 | 1.02 |
| 23 | 24+38 | sFR2 | 327 | 1005 | 297,172 | 6,954,955 | 1.017 |
| 24 | 17+40 | sUK3 | 1032 | 2502 | 452,086 | 7,454,624 | 1.032 |
| 25 | 25+37 | sGER2 | 1399 | 649 | 115,425 | 5,696,016 | 1.008 |
| 26 | 23 | sIT3 | 1715 | 1075 | 519,328 | 7,087,774 | 1.013 |
| 27 | 41 | sNL4 | 596 | 533 | 1,462,561 | 7,751,551 | 1.01 |
| Total |  |  | 12,587 | 23,503 |  |  | 1.015 |

**Imputation**

The imputation process was executed using Genotype Imputation v1.7.4 via the Michigan Imputation Server (Das et al., 2016). The prephasing of genomic data within each stratum was performed using Eagle v2.4, with HapMap2 (GRCh37/hg19) haplotypes. The reference panel was restricted to the European population.

**Quality controls for the validation dataset**

SNPs were annotated based on dbSNP150 and aligned to the hg19 reference genome. Multi-allelic and palindromic SNPs were excluded, followed by initial quality checks per cohort to remove low-quality SNPs and individuals using PLINK 1.9 (--geno 0.1 and --mind 0.1). Population structure was assessed by projecting HapMap3 principal components using EIGENSOFT 6.1.4, with extreme outliers from European ancestries removed (> 25 SD on PC1-4).

After cohort QC and merging, stringent quality control was performed per stratum to further exclude SNPs and individuals. Criteria included MAF > 0.01, SNP genotyping rate > 0.98, deviation from Hardy-Weinberg equilibrium in controls (P > 1 × 10^-5^), and haplotype-biased missingness (P > 1 × 10^-8^).

Subsequent QC thresholds were applied for individual exclusion: individual missingness > 0.02, |F| > 0.2, gender mismatches, and missing phenotypes. SNPs with differential missingness (P < 1 × 10^-4^) were excluded, and duplicate individuals removed (PI_HAT > 0.8). Outliers from European ancestries, as well as outliers within each stratum, were also excluded.

**Evaluation metrics**

1.**Accuracy**: This fundamental metric represents the ratio of correctly predicted instances to the total instances, offering a holistic measure of the model's overall correctness:

$$Accuracy=\frac{TruePositives+TrueNegatives}{TotalInstances}$$

2.**Precision**: Precision focuses on the accuracy of positive predictions, highlighting the model's ability to avoid false positives:

$$Precision=\frac{TruePositives+FalsePositives}{TruePositives}$$

3.**Recall (Sensitivity)**: Recall, or true positive rate, gauges the model's capability to identify all relevant instances, emphasizing its sensitivity to positive cases:

$$Recall=\frac{TruePositives+FalseNegatives}{TruePositives}$$

4.**F1 Score**: The F1 score, a harmonic mean of precision and recall, provides a balanced assessment of the model's performance, particularly valuable when there is an imbalance between positive and negative instances:

$$F1Score=2\times\frac{Precision\times Recall}{Precision+Recall}$$

5.**AUC (Area Under the ROC Curve)**: The AUC quantifies the model's ability to discriminate between positive and negative instances across different probability thresholds. It is derived from the Receiver Operating Characteristic (ROC) curve and ranges from 0 to 1, with higher values indicating superior discriminatory power.

$$AUC=\int_{0}^{1} TPR({FPR}^{-1}(t))ⅆt$$

$$TPR(TruePositiveRate)=\frac{TruePositives}{TruePositives+FalseNegatives}$$

$$FPR(FalsePositiveRate)=\frac{FalsePositives}{FalsePositives+TrueNegatives}$$

Supplementary Table 3. Number of cases and controls in leave-one-out method and leave-half-out method.

|  | **leave-one-out** | | | | | | **leave-half-out** | | | | | |
| --- | --- | --- | --- | --- | --- | --- | --- | --- | --- | --- | --- | --- |
|  | **Training set** | | | **Test set** | | | **Training set** | | | **Test set** | | |
| **Name** | **Controls** | **Cases** | **Total** | **Controls** | **Cases** | **Total** | **Controls** | **Cases** | **Total** | **Controls** | **Cases** | **Total** |
| sBE | 22,944 | 12,083 | 35,027 | 504 | 504 | 1,008 | 23,251 | 12,335 | 35,586 | 252 | 252 | 504 |
| sFIN | 23,028 | 12,071 | 35,099 | 475 | 475 | 950 | 23,266 | 12,350 | 35,616 | 237 | 237 | 474 |
| sFR | 21,844 | 12,105 | 33,949 | 482 | 482 | 964 | 23,262 | 12,346 | 35,608 | 241 | 241 | 482 |
| sGER | 22,596 | 10,670 | 33,266 | 907 | 907 | 1,814 | 23,050 | 12,134 | 35,184 | 453 | 453 | 906 |
| sIB | 23,404 | 12,461 | 35,865 | 99 | 99 | 198 | 23,454 | 12,538 | 35,992 | 49 | 49 | 98 |
| sIR | 22,729 | 12,015 | 34,744 | 572 | 572 | 1,144 | 23,217 | 12,301 | 35,518 | 286 | 286 | 572 |
| sIT | 22,091 | 10,199 | 32,290 | 1,412 | 1,412 | 2,824 | 22,797 | 11,881 | 34,678 | 706 | 706 | 1,412 |
| sNL | 15,823 | 10,471 | 26,294 | 2,116 | 2,116 | 4,232 | 22,445 | 11,529 | 33,974 | 1,058 | 1,058 | 2,116 |
| sSWE | 22,989 | 12,061 | 35,050 | 514 | 514 | 1,028 | 23,246 | 12,330 | 35,576 | 257 | 257 | 514 |
| sSWISS | 23,282 | 12,384 | 35,666 | 203 | 203 | 406 | 23,402 | 12,486 | 35,888 | 101 | 101 | 202 |
| sUK | 18,155 | 10,773 | 28,928 | 1,814 | 1,814 | 3,628 | 22,596 | 11,680 | 34,276 | 907 | 907 | 1,814 |
| sUS | 19,648 | 11,164 | 30,812 | 1,423 | 1,423 | 2,846 | 22,792 | 11,876 | 34,668 | 711 | 711 | 1,422 |

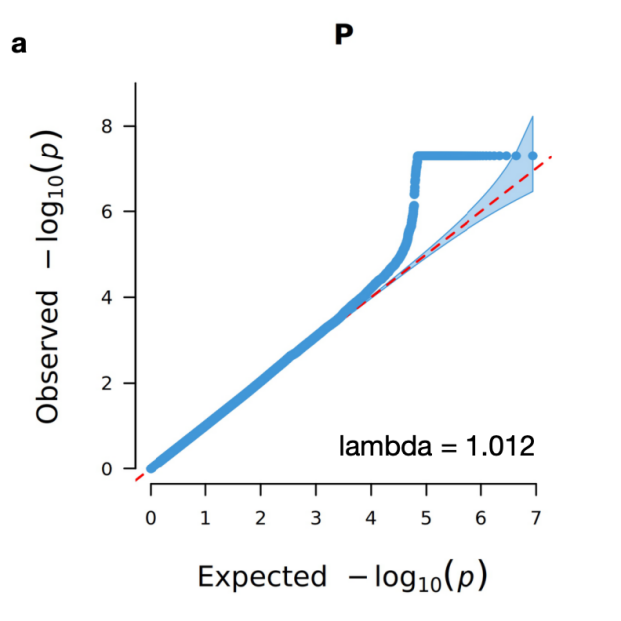

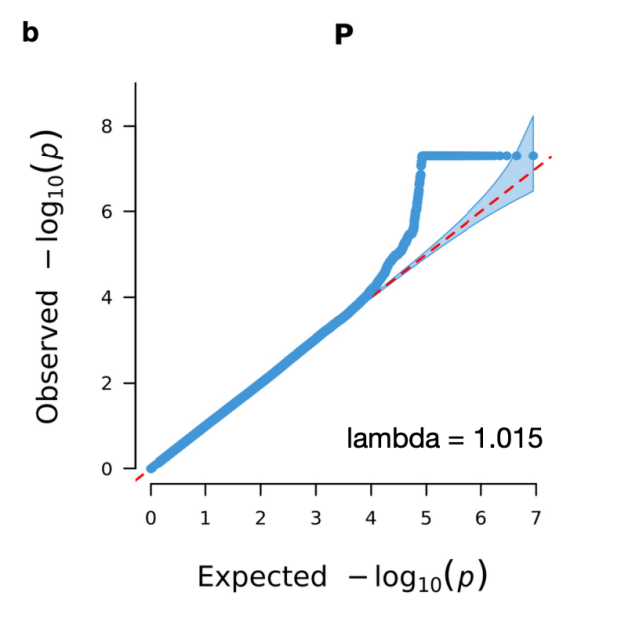

Supplementary Figure 1. Quantile-quantile plot for the meta-analysis results a) of the 2016 published GWAS paper, and of b) our study.

### Hyperparameters

To optimise the DiseaseCapsule model, all hyperparameters were fine-tuned to enhance its training dynamics. Employing the Adam optimization algorithm, all hyperparameters were chosen to facilitate efficient convergence and robust generalization. The initial learning rate, set at 0.0001, served as the foundational step size for model adaptation. To balance rapid convergence with sustained learning adaptability, a decay factor (γ) of 0.8 was employed, dynamically adjusting the learning rate in each epoch through an exponential scheduler. The hyperparameter optimization process involved 50 epochs, where each epoch represented a full pass through the entire dataset. To prevent overfitting, an early stop mechanism was implemented. Specifically, if the model's performance did not show improvement over a continuous span of three consecutive epochs, the optimization process was halted. A batch size of 128 was strategically selected for training iterations.

**References**

Das, S., Forer, L., Schönherr, S., Sidore, C., Locke, A. E., Kwong, A.,…McGue, M. (2016). Next-generation genotype imputation service and methods. *Nature genetics*, *48*(10), 1284-1287.
